# Cohort-Defined Low Skeletal Muscle Mass and Functional Dependency After Critical Illness: A 12-Month Longitudinal Study

**DOI:** 10.64898/2026.09.04.26362259

**Authors:** Billal Gomaa, Ruth-Ann Brown, Anthony S Bonavia

## Abstract

**Objective:** To evaluate the association between low skeletal muscle mass at intensive care unit (ICU) enrollment and functional dependency at 1, 3, 6, and 12 months after enrollment.

**Design:** Prospective longitudinal observational cohort with follow-up at 1, 3, 6, and 12 months after enrollment.

**Exposure:** Sex-defined low muscle mass (LMM), using L3 skeletal muscle index (SMI) thresholds of <33.2 cm^2^/m^2^ in men and <28.3 cm^2^/m^2^ in women, derived from the cohort distribution.

**Main outcome and analysis:** Need for assistance with activities of daily living, transfers, or toileting. Logistic generalized estimating equations (GEE) included LMM-by-time interactions, adjustment for sepsis, sex, and age <65 years, and mortality-derived inverse probability weights.

**Results:** Among 217 participants, 148 had SMI measurements and 15 had sex-defined LMM. The functional GEE included 404 assessments from 121 patients, including 12 with LMM. Observed dependency at 1 month was 8/12 (66.7%) with LMM versus 35/108 (32.4%) without LMM. The adjusted 1-month odds ratio (OR) was 3.52 (95% confidence interval [CI], 0.99-12.44; p=0.051); the 12-month OR was 1.67 (95% CI, 0.41-6.81; p=0.475). The joint LMM-by-time test gave p=0.076. A model imposing a common effect across visits gave OR 2.04 (95% CI, 0.79-5.24). Sensitivity analyses were imprecise and did not establish a persistent association.

**Conclusions:** Low muscle mass may identify patients with greater early functional dependency after critical illness. Sparse exposed numbers, mortality, and incomplete follow-up limit inference about persistence and recovery trajectories. The data do not establish equivalent recovery patterns or a causal effect of muscle mass.

## Introduction

Post-Intensive Care Syndrome (PICS) encompasses physical, cognitive, and mental health impairments persisting after critical illness [1, 2]. Functional dependency, including the need for assistance with activities of daily living (ADLs), transfers, or toileting, is an important patient-centered outcome with implications for quality of life, caregiver burden, and post-discharge care [3, 4].

Skeletal muscle contributes to the physiologic response to critical illness and is vulnerable to catabolism during intensive care [5, 6]. Low skeletal muscle mass (LMM) at enrollment may reflect pre-existing depletion, acute illness-related change, or both. Previous studies have examined its association with mortality and longer-term outcomes, but muscle quantity alone does not establish premorbid functional reserve or the mechanism of subsequent disability [7, 8].

L3 skeletal muscle index (SMI), derived from clinically obtained computed tomography (CT), allows muscle mass to be quantified without additional imaging [9]. Body composition can reveal low muscle quantity that is not apparent from body mass index alone [10]. Associations with mortality have been reported, but the relationship between baseline muscle measurements and later dependency may depend on patient selection, the outcome assessed, and handling of death and incomplete follow-up [11, 12].

This study evaluates the association between cohort-defined LMM and functional dependency at four follow-up visits over 12 months. We distinguish time-specific associations from a common effect across visits and report exploratory analyses addressing exposure definition, continuous SMI, and the combined burden of death or dependency.

## Methods

### Study Design and Patient Population

We analyzed a prospective longitudinal cohort of 217 critically ill patients assessed at 1, 3, 6, and 12 months after enrollment. Critical illness was defined as the need for intravenous vasopressors therapy or non-invasive/invasive respiratory support and delivered in an intensive care unit (ICU) setting. Sepsis status was defined using Sepsis-3 consensus criteria [13]. CT scans were eligible for L3 SMI calculation if they were performed within 10 days of ICU admission and were available for 148 participants. SMI was calculated using the method described by Gomez-Perez et al, and measured using ImageJ 1.54g software [9]. Participants without SMI were retained in baseline and missingness descriptions but excluded from exposure-based models. The number of participants contributing functional outcomes at each visit was determined separately, based on visit-level eligibility and outcome availability.

The primary exposure was sex-defined LMM using cohort-derived thresholds of <33.2 cm^2^/m^2^ for men and <28.3 cm^2^/m^2^ for women. The full-precision sex-specific 10th percentiles were 33.16 and 28.26 cm^2^/m^2^; rounding did not change classification. An age-based sensitivity definition used the full-precision within-age 10th percentiles (33.90 cm^2^/m^2^ for age <65 years and 28.85 cm^2^/m^2^ for age ≥65 years), with SMI at or below the percentile classified as low (Supplementary Figure S1). Classification sensitivity to cutoff precision was assessed by applying strict age thresholds rounded to one decimal place (Supplementary Table S2).

### Outcomes

The primary outcome was functional dependency at each visit, defined as the need for help with at least one of three activities: ADLs, transfers, or toileting. These were assessed by phone calls to the patient or primary caregiver at the pre-defined time points. The questions and scoring method are outlined in the Supplement. A positive component defined dependency; all three components had to be negative to define independence. Otherwise, the composite was missing. In this dataset, the three components were either all observed or all missing; no partially observed component set required additional handling (Supplementary Table S1).

Secondary outcomes were described as observed adverse proportions with outcome-specific denominators: walking, dressing, and stair assistance; mobility limitation; poor performance status; and non-home residence (Supplementary Figure S2). Constructed domain scores and multiple secondary models were exploratory and were not used to support confirmatory conclusions.

### Eligibility, Censoring, and Missingness

Nominal visits corresponded to days 30, 90, 180, and 365. A visit was eligible for functional analysis when no death was recorded before its nominal day, and the visit month was strictly earlier than the recorded censoring month. A missing censoring month was treated as follow-up through 12 months. The data did not separately identify administrative censoring and loss to follow-up, and a missing death date was treated as no documented death. These assumptions were retained for reproducibility and limit interpretation of survivor outcomes.

Recorded outcomes at nominal visits after a documented death or at or after the censoring month were excluded. Missing eligible functional responses were not imputed, and the mortality weights did not model response missingness. We report enrolled, measured-SMI, eligible, and observed-outcome denominators separately. In an exploratory death-or-dependency endpoint, documented death before a visit was adverse; living participants required an observed functional outcome; visits at or after the censoring month remained excluded.

### Statistical Analysis

The primary longitudinal model was a logistic GEE with exchangeable working correlation and patient-clustered robust covariance. Predictors were sex-defined LMM, indicators for 3, 6, and 12 months (reference: 1 month), LMM-by-time interactions, sepsis, sex, and age <65 years. Mortality-derived inverse probability weights were estimated from a Cox model containing LMM and the same adjustment variables among the 148 participants with SMI. A recorded death date defined an event; participants without a recorded death date were assigned follow-up through day 365. At each eligible visit the weight was the inverse estimated probability of surviving to that nominal day.

The LMM main coefficient estimates the adjusted contrast at 1 month. Later visit contrasts add the respective interaction coefficient and use its covariance with the main coefficient. We report 95% Wald confidence intervals and a joint three-degree-of-freedom Wald test of all LMM-by-time terms. Weighted predictions were standardized over the baseline covariate profiles of all 148 measured-SMI participants; pointwise intervals used the delta method and treated the standardization covariates as fixed.

Exploratory sensitivity analyses included unweighted exchangeable GEE, models imposing a common effect across visits, independent-working-correlation GEE among eligible survivors with ordinary and bias-reduced robust covariance, age-based LMM, and continuous SMI per 10 cm^2^/m^2^ lower. Independent working correlation was examined because partly conditional survivor means require care when observations are truncated by death [14, 15]. Continuous-SMI models used a common linear log-odds slope and an exploratory three-knot restricted cubic spline to avoid relying solely on dichotomization [16]. A separate independent-working-correlation model examined death or dependency. These sensitivity models address different assumptions or endpoints and were not selected on their p-values.

In a post hoc sensitivity analysis, we added a single indicator for documented peri-enrollment hydrocortisone use to the primary logistic GEE. Explicit positive doses defined documented use; explicit no-use statements or zero-dose schedules defined no documented use, and N/A entries were treated as missing. The original covariates, LMM-by-time interactions, exchangeable correlation, robust patient-clustered covariance, eligibility rules, and mortality weights were retained. We compared models with and without the hydrocortisone indicator in the same participants with known steroid status.

Additional checks comprised empirical-cutoff bootstrap intervals, leave-one-exposed-patient-out refits of the Cox model and GEE, and 1,000 patient-level bootstrap refits retaining the original sex-defined labels. Numerical failures and extreme coefficients were retained as diagnostics. Exploratory analyses were not adjusted for multiplicity, and interpretation emphasized uncertainty. Analyses used Python 3.8.18, *statsmodels* 0.14.0, and *lifelines* 0.27.8. Detailed weighting and reproducibility information is provided in the Supplement.

## Results

### Cohort Characteristics and Exposure Availability

The dataset contained 217 unique participants, including 148 with SMI: 133 without sex-defined LMM and 15 with LMM (Table 1). SMI was unavailable in 69/217 (31.8%). Availability differed by sepsis status: 114/145 (78.6%) among participants with sepsis versus 34/72 (47.2%) without sepsis. The primary functional GEE included 404 assessments from 121 participants, including 12 with LMM.

**Table 1.** Baseline characteristics by CT-based skeletal muscle mass availability and sex-defined low muscle mass status.

| Characteristic | Overall | No sex-defined LMM | Sex-defined LMM | SMI unavailable |
| --- | --- | --- | --- | --- |
| Patients, n | 217 | 133 | 15 | 69 |
| Male sex, n/N (%) | 116/217 (53.5%) | 72/133 (54.1%) | 8/15 (53.3%) | 36/69 (52.2%) |
| Age <65 years, n/N (%) | 87/217 (40.1%) | 51/133 (38.3%) | 1/15 (6.7%) | 35/69 (50.7%) |
| Sepsis, n/N (%) | 145/217 (66.8%) | 102/133 (76.7%) | 12/15 (80.0%) | 31/69 (44.9%) |
| In-hospital mortality, n/N (%) | 30/217 (13.8%) | 19/133 (14.3%) | 2/15 (13.3%) | 9/69 (13.0%) |
| Recorded death date, n/N (%) | 73/217 (33.6%) | 44/133 (33.1%) | 6/15 (40.0%) | 23/69 (33.3%) |
| Recorded death before day 30, n/N (%) | 36/217 (16.6%) | 23/133 (17.3%) | 3/15 (20.0%) | 10/69 (14.5%) |
| L3 SMI, cm<sup>2</sup>/m<sup>2</sup>, median (IQR) | 43.0 (35.4-53.3) | 44.8 (38.2-54.8) | 27.0 (24.9-30.1) | NA |

LMM, low muscle mass; SMI, skeletal muscle index. Sex-defined LMM uses <33.2 cm<sup>2</sup>/m<sup>2</sup> for men and <28.3 cm<sup>2</sup>/m<sup>2</sup> for women. Recorded death-date counts describe available records; missing dates do not establish survival. One participant with unavailable SMI had in-hospital mortality recorded without a death date.

### Follow-up Eligibility and Primary Outcome

Within the measured-SMI cohort, 122, 110, 94, and 89 participants were eligible at 1, 3, 6, and 12 months; 120, 105, 93, and 86 had observed primary outcomes, respectively (Table 2). Eleven eligible assessments lacked all three components, all in the no-LMM group. Observed proportions and weighted standardized estimates are shown in Figure 1. Full visit-level counts are provided in Supplementary Table S5.

**Figure 1.**
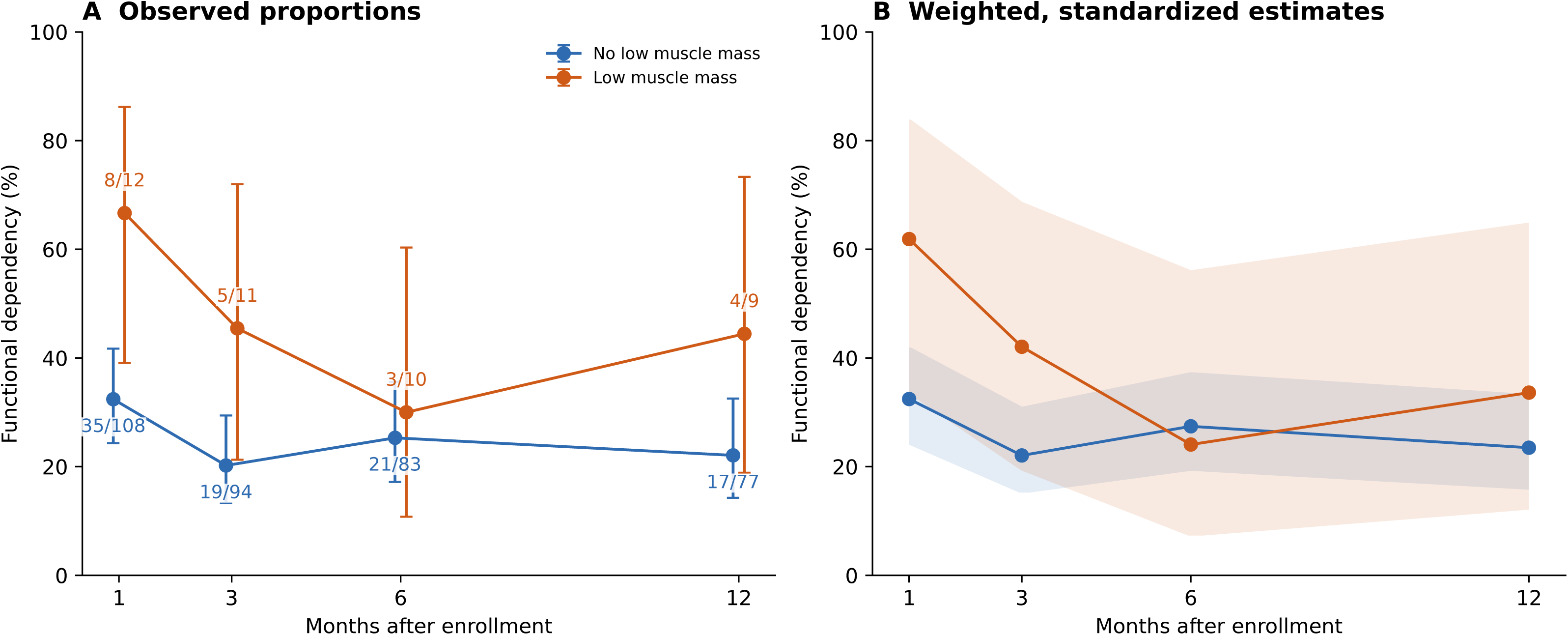
Observed and weighted standardized functional dependency over follow-up. Panel A: observed proportions among eligible respondents, with Wilson 95% confidence intervals and dependent/observed counts. Panel B: predictions from the mortality-weighted interaction GEE, standardized over baseline covariates of the 148 participants with SMI; shading denotes pointwise 95% delta-method intervals. These intervals do not incorporate uncertainty from estimated mortality weights or cohort-derived cutoffs. Lines connect nominal visits for display and do not represent individual recovery trajectories.

**Table 2.** Visit eligibility and observed functional dependency in participants with measured SMI.

| Follow-up | Eligible with SMI, n | No LMM: dependent/observed | LMM: dependent/observed | Eligible with missing outcome, n |
| --- | --- | --- | --- | --- |
| 1 month | 122 | 35/108 (32.4%) | 8/12 (66.7%) | 2 |
| 3 months | 110 | 19/94 (20.2%) | 5/11 (45.5%) | 5 |
| 6 months | 94 | 21/83 (25.3%) | 3/10 (30.0%) | 1 |
| 12 months | 89 | 17/77 (22.1%) | 4/9 (44.4%) | 3 |

Cells for functional dependency show dependent/observed n/N (%). Eligible counts use recorded death dates and censoring months. Missing eligible outcomes are excluded from the observed denominators; all 11 missing assessments occurred in the no-LMM group.

Observed dependency was 8/12 (66.7%) versus 35/108 (32.4%) at 1 month, 5/11 (45.5%) versus 19/94 (20.2%) at 3 months, 3/10 (30.0%) versus 21/83 (25.3%) at 6 months, and 4/9 (44.4%) versus 17/77 (22.1%) at 12 months (LMM versus no LMM). These proportions describe changing sets of eligible respondents, not individual recovery slopes.

### Primary Longitudinal Association and Sensitivity Checks

The weighted adjusted LMM odds ratio at 1 month was 3.52 (95% CI, 0.99-12.44; p=0.051). The 3-, 6-, and 12-month contrasts were 2.63 (0.78-8.88), 0.84 (0.19-3.73), and 1.67 (0.41-6.81), respectively (Table 3; Figure 2). The joint LMM-by-time test gave χ²=6.89 with 3 degrees of freedom (p=0.076). Estimated associations were imprecise, and the interaction result did not establish either differential or equivalent recovery patterns.

**Figure 2.**
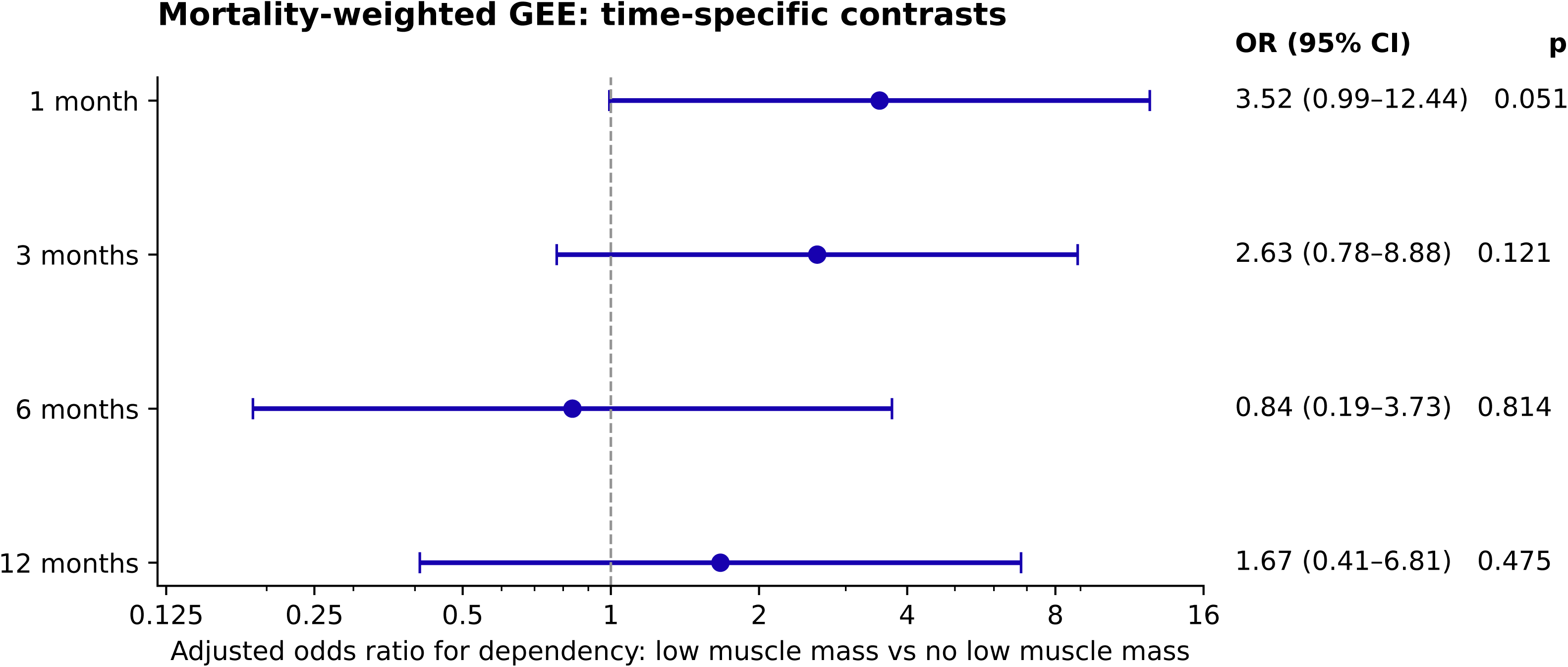
Time-specific low-muscle-mass contrasts from the mortality-weighted GEE. Points show adjusted odds ratios comparing sex-defined LMM with no LMM at each nominal visit; bars show 95% Wald confidence intervals on a logarithmic scale. The vertical line marks OR=1. Models adjusted for sepsis, sex, and age <65 years and included LMM-by-time interactions. The 1-month contrast is the main LMM coefficient; subsequent contrasts combine it with the relevant interaction. The analysis included 404 assessments from 121 patients, 12 with LMM.

**Table 3.** Time-specific sex-defined LMM contrasts from the mortality-weighted interaction GEE.

| Follow-up | Adjusted OR | 95% CI | p |
| --- | --- | --- | --- |
| 1 month | 3.52 | 0.99-12.44 | 0.051 |
| 3 months | 2.63 | 0.78-8.88 | 0.121 |
| 6 months | 0.84 | 0.19-3.73 | 0.814 |
| 12 months | 1.67 | 0.41-6.81 | 0.475 |

OR, odds ratio; CI, confidence interval. The model included 404 assessments from 121 patients and adjusted for sepsis, sex, and age <65 years. Each row compares LMM with no LMM at that visit. The joint LMM-by-time Wald test gave $\chi^2=6.89$ (3 df), $p=0.076$ . These are not estimates of a common year-long effect.

The unweighted exchangeable interaction model gave a 1-month OR of 3.54 (95% CI, 1.02-12.27; p=0.046). A weighted common-effect model gave OR 2.04 (0.79-5.24; p=0.139). Independent-working-correlation GEE with bias-reduced covariance gave a 1-month OR of 3.54 (0.90-13.97; p=0.071). The continuous-SMI common-slope model gave OR 1.23 (0.89-1.71; p=0.216) per 10 cm^2^/m^2^ lower SMI. The full-precision age-based definition classified 16 participants as LMM, including 12 with functional outcomes, and gave a 1-month OR of 2.22 (0.68-7.17; p=0.184). The death-or-dependency model included 559 observations from 147 patients and gave a common OR of 1.86 (0.76-4.55; p=0.174). Supplementary Tables S2-S4 provide definitions and additional estimates.

In a post hoc sensitivity analysis of 112 participants with known hydrocortisone status (381 assessments), the 1-month LMM odds ratio changed from 3.24 (95% CI 0.91-11.54) to 2.89 (95% CI 0.83-10.12) after adjustment for documented use (p=0.096). This analysis retained all 12 participants with LMM and the original model specification and mortality weights.

## Discussion

In this cohort, sex-defined LMM was associated with higher estimated odds of dependency at 1 month, but uncertainty was substantial. Later contrasts and common-effect sensitivity models were also imprecise. The findings support a possible early association requiring confirmation; they do not establish a persistent upward shift in disability, a causal role for muscle mass, or equivalence of recovery patterns.

A lower-reserve phenotype is a plausible hypothesis, but it was not directly measured here. Premorbid function was unavailable, and baseline SMI alone cannot distinguish pre-existing depletion from acute illness-related change. Group proportions across visits also reflect death, censoring, and changing respondent composition. Consequently, any pattern in these proportions cannot establish a biological recovery slope. Prior sepsis studies motivate investigation of muscle mass and longer-term outcomes [7, 8]. The TRACER publication, however, is a prospective study protocol rather than evidence validating equal recovery trajectories [17].

The BRAIN-ICU secondary analysis by Rengel and colleagues evaluated muscle characteristics, disability, and self-reported physical function in critical-illness survivors [18]. Differences between studies may reflect exposure measurement, outcomes, selection, and incomplete follow-up. Our results do not demonstrate that survival bias explains another study’s findings. Inverse probability methods can address some forms of dependent censoring under appropriate assumptions [19, 20], but death also changes the meaning of a functional outcome. Survivor-conditional, death-inclusive, and hypothetical mortality-weighted questions should be distinguished [14, 15]. The weighting used here depends on the recorded mortality information and the specified Cox model; it neither imputes a functional state after death nor establishes removal of survivor bias.

The sex- and age-based definitions identify small, partly different groups. Their p-values do not test comparative predictive sensitivity, discrimination, calibration, or clinical utility. Sex-specific muscle distributions provide a rationale for stratification [21], and sex-related muscle biology may be relevant in critical illness [22, 23], but neither establishes the performance of this cohort’s thresholds. The classification change produced by rounding an age cutoff illustrates the need for a shared full-precision exposure definition. Continuous SMI analyses and external validation should complement categorical descriptions [24].

Opportunistic CT-based muscle assessment remains a candidate prognostic measure, and artificial intelligence-powered tools may facilitate its measurement when suitable clinical imaging is available. This study does not establish a screening threshold, a treatment-selection rule, or benefit from interventions directed specifically at low SMI. Functional assessment, nutrition, rehabilitation, and discharge-support needs should be evaluated in future studies that measure these factors directly.

Several limitations constrain inference. Factors influencing muscle mass and physical conditioning before and during critical illness were incompletely characterized, including premorbid function, frailty, habitual physical activity, chronic corticosteroid exposure, and nutritional status. We also lacked sufficient information to account for energy and protein delivery, mobilization, and the timing, intensity, and duration of physical therapy during hospitalization and recovery. Although a sensitivity analysis adjusted for documented peri-enrollment hydrocortisone use, it did not comprehensively capture chronic corticosteroid use, other glucocorticoids, cumulative exposure, or treatment timing relative to CT. Residual confounding by these factors, illness severity, and muscle quality limits attribution of subsequent dependency specifically to low muscle mass. SMI was missing in nearly one-third of participants and was more frequently available among those with sepsis, creating potential selection bias.

Only 12 participants with LMM contributed functional outcomes, with 9-12 respondents per visit, limiting precision and the scope for additional covariate adjustment. The dataset did not distinguish administrative censoring from dropout, and complete vital-status ascertainment and actual assessment timing could not be independently verified. Responses at nominal visits after recorded death or at or after censoring were excluded. Mortality weights did not address missing functional responses or missing SMI, and conventional robust covariance treated estimated weights as fixed.

## Conclusion

Low muscle mass may identify patients with greater early functional dependency after critical illness, but the association was imprecisely estimated in this small exposed subgroup. The available data do not establish persistence, equivalent recovery rates, or superiority of sex-defined thresholds. Further study with more complete ascertainment, premorbid functional measurements, and external validation is needed before clinical prediction or treatment-selection claims can be made.

## Declarations

### Ethics Approval and Consent to Participate

This study was approved by the Penn State Human Subjects Protection Office (#15328, approved 8/6/2020). All participants and/or their legally authorized representatives provided informed consent to participate.

### Author Contributions

Billal Gomaa: Investigation, Data curation, Visualization, Writing - original draft, Writing - review and editing. Ruth-Ann Brown: Investigation, Data curation, Writing – review and editing. Anthony Bonavia: Conceptualization, Methodology, Data curation. Formal analysis, Software, Validation, Visualization, Supervision, Project administration, Funding acquisition, Writing – original draft, Writing – reviewing and editing. All authors reviewed and approved the final manuscript and agree to be accountable for the work.

### Declaration of AI-assisted technologies

During the preparation of this work, the authors used OpenAI Codex to assist with analysis and statistical code, editorial review, identification of reporting and formatting issues, and drafting submission materials. The authors reviewed and verified all AI-assisted material and analytical outputs, checked the accuracy of AI-assisted text, and edited the content as needed. The authors take full responsibility for the analyses, interpretation, and content of the published article.

## Data Availability

All data produced in the present study are available upon reasonable request to the authors

