## Supplement for "Cohort-Defined Low Skeletal Muscle Mass and Functional Dependency After Critical Illness: A 12-Month Longitudinal Study"

Supplementary methods for the cohort analysis — p.2

Outcome availability and follow-up exclusions — p.2

Influence and bootstrap analyses — p.2

Table S1. Definitions of primary and descriptive outcomes — p.3

Figure S1. SMI distributions and empirical cutoffs — p.4

Table S2. Empirical SMI cutoffs and age-based sensitivity analysis — p.4

Figure S2. Observed adverse outcomes among eligible respondents — p.5

Table S3. Primary and sensitivity model estimates — p.6

Table S4. Death-or-dependency endpoint — p.6

Table S5. Follow-up eligibility and outcome availability — p.7

Functional dependency questionnaire and scoring — p.8

Questionnaire items and outcome coding — p.8

### Supplementary methods for the cohort analysis

#### Outcome and follow-up eligibility

Functional dependency was defined from reported assistance with activities of daily living (ADLs), transfers, or toileting; the questionnaire wording and scoring appear below. Follow-up visits were evaluated at nominal days 30, 90, 180, and 365. A visit was eligible when no death was recorded before its nominal day and the visit preceded the recorded censoring month. Participants with no recorded censoring month were considered eligible through day 365. The data did not distinguish administrative censoring from loss to follow-up.

#### Primary longitudinal model

The primary logistic generalized estimating equation (GEE) used an exchangeable working correlation and patient-clustered robust covariance. The model included categorical time, with 1 month as the reference; low muscle mass (LMM); LMM-by-time interactions; sepsis; sex; and age younger than 65 years. The LMM coefficient estimates the 1-month contrast, and each later contrast combines that coefficient with the corresponding interaction term. The joint interaction test had 3 degrees of freedom. Adjusted estimates were standardized over the baseline covariate distribution of the 148 participants with measured skeletal muscle index (SMI).

#### Mortality weighting

Mortality weights were estimated with a Cox model among the 148 participants with measured SMI. The model included LMM, sepsis, sex, and age younger than 65 years. Recorded death dates defined events; participants without a recorded death date contributed event-free follow-up through day 365. At each eligible visit, the weight was the inverse estimated probability of survival to that nominal day. Weights ranged from 1.168 to 1.658 (median, 1.369; 99th percentile, 1.607). A numerical survival-probability floor of 10⁻⁶ was specified but was not reached. Standard errors treated the estimated weights as fixed.

#### Sensitivity analyses

Sensitivity analyses included unweighted GEE models, common-effect models without LMM-by-time interactions, independent-working-correlation models with ordinary or bias-reduced robust covariance, an age-based LMM definition, continuous SMI per 10 cm^2^/m^2^ lower, and a three-knot restricted cubic spline using the 10th, 50th, and 90th percentiles of SMI. A separate model evaluated death or dependency. These analyses were exploratory, and no adjustment was made for multiple comparisons.

### Outcome availability and follow-up exclusions

Eleven eligible assessments lacked all three primary outcome components; all occurred in participants without LMM. Table S5 presents bounds that classify these responses as either independent or dependent. Seventeen responses from 14 participants occurred at nominal visits after a recorded death, and four occurred at or after the recorded censoring month; these observations were excluded. Two participants with measured SMI had recorded day-zero deaths. One participant without SMI had recorded in-hospital mortality without a death date. Exact assessment dates, reasons for censoring, and complete vital status were unavailable.

### Influence and bootstrap analyses

Leave-one-participant-out analyses among the 12 participants with LMM yielded 1-month odds ratios (ORs) of 3.04-4.72 and p values of 0.026-0.088. A seeded patient-level bootstrap resampled all 148 participants with measured SMI 1,000 times, held the sex-specific LMM classifications fixed, and refitted the Cox and GEE models. Finite estimates were obtained in 960 samples; 51 of these had an absolute time-specific log OR greater than 20, indicating severe numerical instability. The descriptive 1-month percentile interval was approximately 0.97-17.24 and did not incorporate cutoff-selection uncertainty. Bias-reduced covariance was evaluated only in unweighted models because it was unavailable for weighted GEE in *statsmodels* 0.14.0. Analyses used Python 3.8.18, *statsmodels* 0.14.0, and *lifelines* 0.27.8.

**Table S1. Definitions of primary and descriptive outcomes**

| **Outcome** | **Components** | **Direction / transformation** | **Combination and missingness** |
| --- | --- | --- | --- |
| Functional dependency | ADL, transfer, toileting assistance | Each component: 1 indicates assistance | Any positive 🡪 positive; all three negative 🡪 negative; otherwise missing. No partial-component rows occurred. |
| Individual assistance outcomes | ADL, transfer, toileting, walking, dressing, and stair assistance | Assistance reported is adverse | Each item is reported separately with an outcome-specific denominator. |
| Mobility limitation (descriptive) | Raw mobility categories 0-3 | Adverse: bed-bound, chair-bound, or assisted mobility; raw 0-2 | Independent mobility (raw 3) is non-adverse. |
| Poor performance (descriptive) | Zubrod/ECOG categories | Adverse: score ≥2 among eligible survivors | Unknown/unavailable scores excluded from outcome denominator. |
| Non-home residence (descriptive) | Residence code | Adverse: assisted living or hospital; home is non-adverse | Unknown residence excluded from denominator. |

ADL, activity of daily living; ECOG, Eastern Cooperative Oncology Group; LMM, low muscle mass.

**Figure S1. SMI distributions and empirical cutoffs**

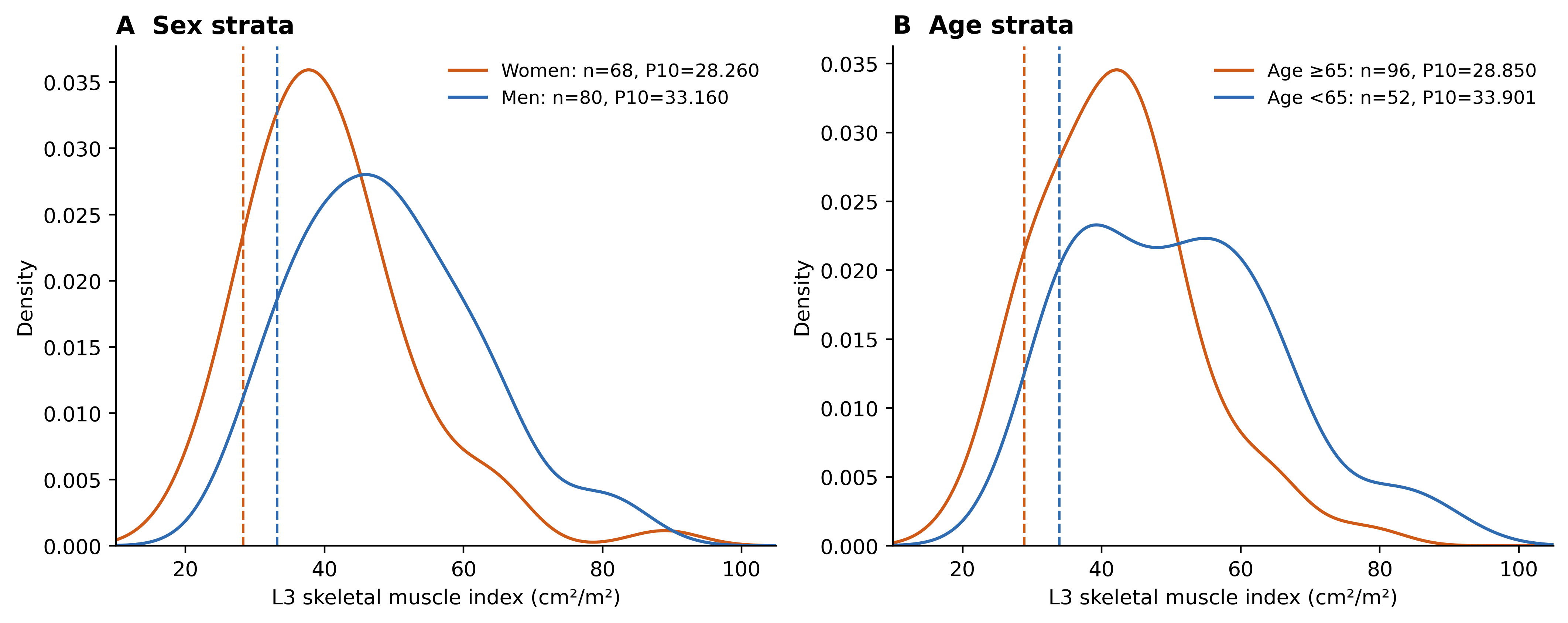

Kernel density estimates show SMI among 80 men, 68 women, 52 participants younger than 65 years, and 96 participants aged 65 years or older. Dashed lines mark empirical 10th percentiles. The primary sex-specific thresholds were <33.2 cm^2^/m^2^ for men and <28.3 cm^2^/m^2^ for women; rounding did not alter classification. The age-based sensitivity thresholds were 33.901 cm^2^/m^2^ for participants younger than 65 years and 28.850 cm^2^/m^2^ for those aged 65 years or older.

**Table S2. Empirical SMI cutoffs and age-based sensitivity analysis**

| **Stratum** | **n** | **SMI P10** | **Bootstrap 95% interval** |
| --- | --- | --- | --- |
| Women | 68 | 28.260 | 24.30-31.04 |
| Men | 80 | 33.160 | 30.19-36.95 |
| Age ≥65 | 96 | 28.850 | 25.98-30.65 |
| Age <65 | 52 | 33.901 | 31.80-37.59 |

Cutoff intervals were estimated from 10,000 within-stratum bootstrap samples (seed, 20260903); SMI is reported in cm^2^/m^2^. The age-based definition classified 16 of 148 participants as having LMM, including 12 with functional follow-up. Its adjusted 1-month OR was 2.22 (95% CI, 0.68-7.17; p = 0.184).

**Figure S2. Observed adverse outcomes among eligible respondents**

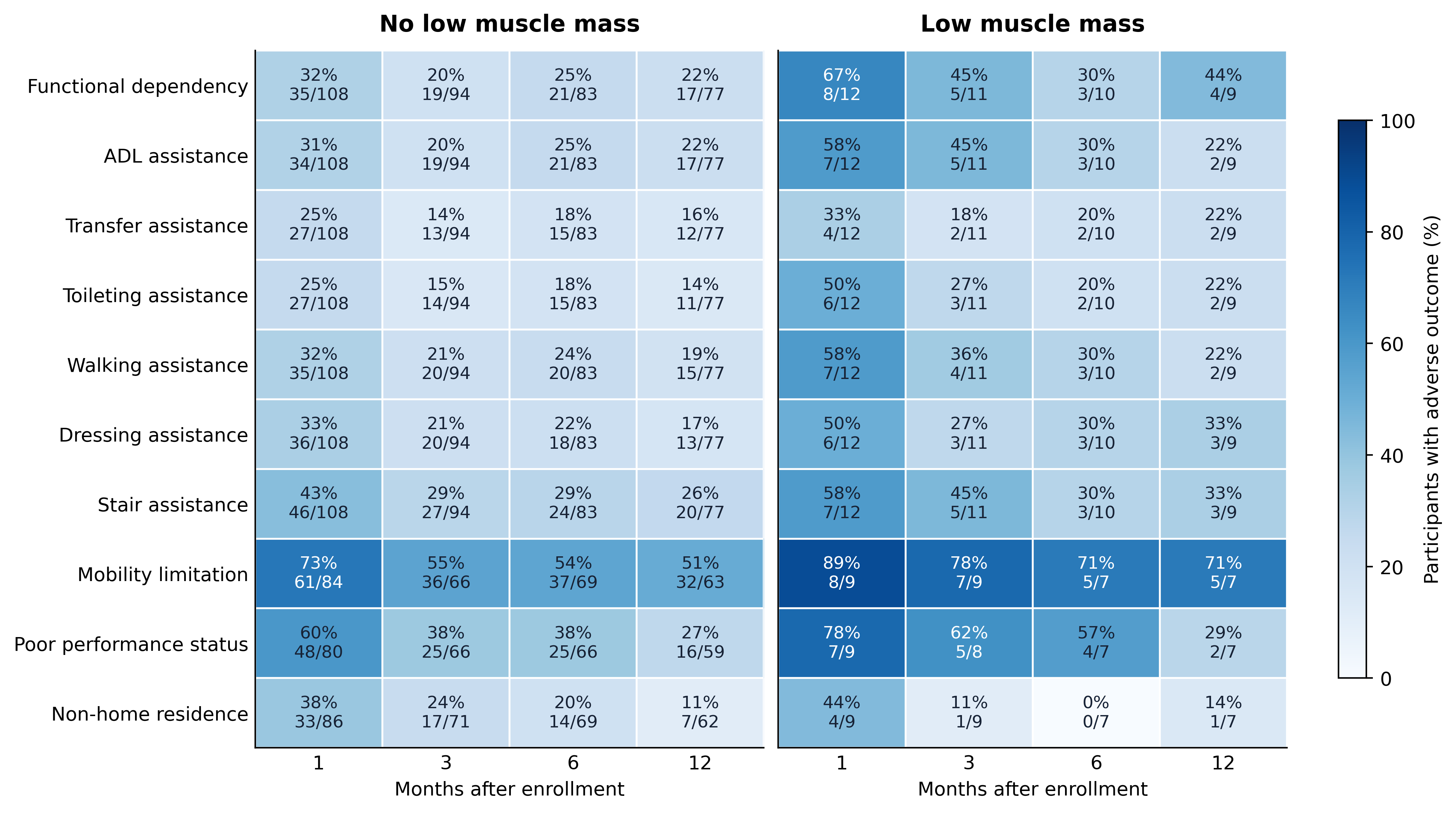

Cells show the percentage with each adverse outcome and the corresponding n/N. Denominators vary by outcome, LMM group, and visit; comparisons are descriptive.

**Table S3. Primary and sensitivity model estimates**

| **Model / contrast** | **Visits / patients** | **OR (95% CI)** | **p** |
| --- | --- | --- | --- |
| Weighted interaction: 1 month | 404 / 121 | 3.52 (0.99-12.44) | 0.051 |
| Unweighted exchangeable interaction: 1 month | 404 / 121 | 3.54 (1.02-12.27) | 0.046 |
| Weighted common effect | 404 / 121 | 2.04 (0.79-5.24) | 0.139 |
| Unweighted common effect | 404 / 121 | 2.12 (0.83-5.41) | 0.116 |
| Independent survivor interaction: 1 month | 404 / 121 | 3.54 (1.00-12.46) | 0.049 |
| Independent survivor interaction: 1 month, bias-reduced SE | 404 / 121 | 3.54 (0.90-13.97) | 0.071 |
| Independent survivor model, common effect | 404 / 121 | 2.25 (0.86-5.90) | 0.097 |
| Continuous SMI: per 10 cm^2^/m^2^ lower | 404 / 121 | 1.23 (0.89-1.71) | 0.216 |
| Death or dependency: independent common effect | 559 / 147 | 1.86 (0.76-4.55) | 0.174 |
| Joint interaction: weighted exchangeable | 404 / 121 | — | 0.076 |
| Joint interaction: unweighted exchangeable | 404 / 121 | — | 0.056 |
| Joint interaction: independent | 404 / 121 | — | 0.366 |
| Joint interaction: independent, bias-reduced SE | 404 / 121 | — | 0.449 |
| Continuous SMI spline: nonlinearity | 404 / 121 | — | 0.384 |
| Continuous SMI spline: overall association | 404 / 121 | — | 0.345 |

Models adjusted for sepsis, sex, and age younger than 65 years. Interaction-model estimates are 1-month contrasts. Common-effect models impose a constant OR across visits. Continuous SMI estimates are per 10 cm^2^/m^2^ lower. Independent-working-correlation models were unweighted. CI, confidence interval; GEE, generalized estimating equation; LMM, low muscle mass; OR, odds ratio; SE, standard error; SMI, skeletal muscle index.

**Table S4. Death-or-dependency endpoint**

| **Follow-up** | **No LMM: adverse/observed** | **LMM: adverse/observed** |
| --- | --- | --- |
| 1 month | 58/131 (44.3%) | 11/15 (73.3%) |
| 3 months | 53/128 (41.4%) | 9/15 (60.0%) |
| 6 months | 59/121 (48.8%) | 8/15 (53.3%) |
| 12 months | 59/119 (49.6%) | 10/15 (66.7%) |

Death recorded before a nominal visit was classified as adverse. Participants without a recorded death required an observed functional outcome. Visits at or after the recorded censoring month were excluded. The model included 559 visits from 147 participants. LMM, low muscle mass.

**Table S5. Follow-up eligibility and outcome availability**

| **Month** | **LMM** | **Baseline n** | **Death before visit** | **At/after censor month** | **Eligible n** | **Observed n** | **Dependent n** | **Missing eligible n** | **Missing-response bounds, %** |
| --- | --- | --- | --- | --- | --- | --- | --- | --- | --- |
| 1 | No | 133 | 23 | 0 | 110 | 108 | 35 | 2 | 31.8-33.6 |
| 1 | Yes | 15 | 3 | 0 | 12 | 12 | 8 | 0 | 66.7-66.7 |
| 3 | No | 133 | 34 | 0 | 99 | 94 | 19 | 5 | 19.2-24.2 |
| 3 | Yes | 15 | 4 | 0 | 11 | 11 | 5 | 0 | 45.5-45.5 |
| 6 | No | 133 | 40 | 11 | 84 | 83 | 21 | 1 | 25.0-26.2 |
| 6 | Yes | 15 | 5 | 0 | 10 | 10 | 3 | 0 | 30.0-30.0 |
| 12 | No | 133 | 44 | 11 | 80 | 77 | 17 | 3 | 21.2-25.0 |
| 12 | Yes | 15 | 6 | 0 | 9 | 9 | 4 | 0 | 44.4-44.4 |

Death and censoring could overlap. Missing-response bounds classify all missing eligible functional responses as either independent or dependent; they do not address missing SMI or incomplete vital-status ascertainment. In the full cohort of 217 participants, 181, 163, 137, and 126 were eligible at 1, 3, 6, and 12 months, respectively. LMM, low muscle mass.

### Functional dependency questionnaire and scoring

Functional dependency was assessed with assistance items from a standardized telephone questionnaire completed by the patient or caregiver. The same questionnaire was administered at 1, 3, 6, and 12 months. Respondents were asked, “Do you need help with any of the following activities?” Three of the six items contributed to the primary outcome.

Questionnaire items and outcome coding

| **Questionnaire item** | **Included in the primary outcome** |
| --- | --- |
| Walking across a small room | No |
| Bathing or dressing yourself | No |
| Transferring from a bed to chair | Yes |
| Using the toilet | Yes |
| Performing your daily living activities | Yes |
| Walking up and down stairs | No |

Each observed component was coded as 1 when assistance was reported and 0 when no assistance was reported. The primary outcome was classified as follows:

- Dependent (1): assistance reported for at least one selected activity.
- Independent (0): no assistance reported for all three selected activities.
- Missing: no positive component and insufficient information to confirm that all three components were negative.

The three selected components were either jointly observed or jointly missing; no assessment contained a partially missing set. The general ADL item was recorded separately and was not calculated from the other questionnaire items.

The composite captures any reported assistance but does not quantify the number or severity of limitations. The separate mobility rating and Zubrod/ECOG performance score were not included in the primary outcome.
